# Evaluation of Bayesian Point-Based System on the Variant Classification of Hereditary Cancer Predisposition Genes

**DOI:** 10.1101/2024.03.04.24303679

**Authors:** Mohammad K. Eldomery, Jamie L. Maciaszek, Taylor Cain, Victor Pastor Loyola, Suraj Sarvode Mothi, David A. Wheeler, Li Tang, Lu Wang, Jeffery M. Klco, Patrick R. Blackburn

## Abstract

**Purpose:** To assess the differences in variant classifications using the ACMG/AMP 2015 guidelines and the Bayesian point-based classification system (here referred to as the point system) in 115 hereditary cancer predisposition genes and explore the utility of the point system in variant sub-tiering.

**Methods:** Germline variant classifications for 721 pediatric patients from an in-house panel were retrospectively evaluated using the two scoring systems.

**Results:** 2376 unique variants were identified. The point system exhibited a lower rate of unique variants of uncertain significance (VUS) of ∼15% compared to ∼36% using the ACMG/AMP 2015 guidelines (p-value < 0.001). This reduction is attributed to the classification of variants as likely benign with one benign supporting evidence (∼12%) or one benign strong evidence (∼4%) using the point system. In addition, the point system resolves conflicting criteria/evidence not recognized by the ACMG/AMP 2015 guidelines (∼5%). Sub-tiering unique VUS calls by the point system indicates ∼11.5% were VUS-Low (0-1 points), while the remaining ∼3.5% were VUS-Mid (2-3 points) and VUS-High (4-5 points).

**Conclusion:** The point system reduces the VUS rate and facilitates sub-tiering. Future large-scale studies are warranted to explore the impact of the point system on improving VUS reporting and/or VUS clinical management.

## INTRODUCTION

Genetic variant curation and analysis is an essential aspect of the practice of genomic medicine. Efforts led by the American College of Medical Genetics and Genomics and the Association for Molecular Pathology (ACMG/AMP) provided a framework for germline variant curation in 2015 (here referred to as ACMG/AMP 2015 guidelines).^1^ Despite the success of this framework, discordance in variant classification between laboratories remained and it was evident that additional clarification and refinement of these guidelines was needed.^2^ Several studies have documented the necessity of periodic and systematic reanalysis of germline variants to improve patient care^3–5^ and to accelerate disease-gene discovery in research settings.^6^

In 2018, the Clinical Genome Resource (ClinGen) Sequence Variant Interpretation (SVI) Working Group (https://clinicalgenome.org/working-groups/sequence-variant-interpretation/) presented a Bayesian classification framework that aims to provide a quantitative approach to variant classification.^7^ Subsequently, Tavtigian and colleagues proposed a model to transform the Bayesian framework to a point-based classification system (here referred to as the point system) to facilitate the integration of the Bayesian framework with the ACMG/AMP 2015 guidelines.^8^ Building on these developments, the objective of our study is to assess the concordance in germline variant classifications using the ACMG/AMP 2015 guidelines and the point system by utilizing data from a cohort of pediatric cancer patients evaluated by a 115-gene cancer predisposition germline panel. We assess the degree of concordance between the two scoring methods and identify discrepancies and potential limitations of each scoring system. Furthermore, we explore the utility of the point system in sub-tiering variants.

## MATERIALS AND METHODS

### Samples and data collection

Variant analysis was performed on germline variant calls from 721 patients who underwent hereditary cancer panel testing between June 2021 and May 2023. Dual Genome Sequencing (GS) and Exome Sequencing (ES) were performed on germline samples (peripheral blood or skin biopsy) to assess for single nucleotide variants, insertions, deletions, and copy number variants in 115 genes associated with the risk of inherited cancer syndromes (see Supplementary Table 1 for complete gene list and preferred transcripts). Sequencing and bioinformatics analysis were performed as previously described.^9,10^

Within our cohort, most evidence codes were applied in accordance with the strength originally proposed in the ACMG/AMP 2015 guidelines. However, a subset of variants had modified strengths either based on our committee review or recommendations from the ClinGen SVI Working Group. In order to make direct comparisons, results were split into variants with the original strengths according to ACMG/AMP 2015 guidelines and variants with modified strengths. The ClinGen-modified evidence codes include: PVS1, PS2/PM6, PS3/BS3, PM2, PM3, and PP3/BP4. Since the BA1-stand-alone evidence code was not included in the point system and biocurators do not perform a comprehensive evaluation for variants meeting BA1, variants meeting the criteria for BA1-stand-alone were excluded.^7^

### Dataset assembly and scoring methodologies

To assemble our dataset, we queried our internal clinical database and retrieved variants classified by the Molecular Pathology/Clinical Genomics laboratory at St. Jude Children’s Research Hospital between June 2021 and May 2023. The corresponding evidence codes and strength of each variant were scored using the ACMG/AMP 2015 guidelines and the point system. Our analysis did not include copy number variations.

### Statistical analysis

Cohen’s kappa statistic was used to assess pairwise concordance between the variant classification generated by the ACMG/AMP 2015 guidelines and the point system. The variant classification was binned by comparing variants of uncertain significance (VUS) across the ACMG/AMP 2015 and the point system against the remaining categories (Benign ‘B’, Likely Benign ‘LB’, Likely Pathogenic ‘LP’, or Pathogenic ‘P’) grouped into a single category labeled as ‘other categories’. This facilitated the utilization of two-by-two tables for testing classification differences using the chi-squared (χ^2^) test. All statistical analyses used a two-sided alpha level of 0.05 and were performed using the R software (version 4.3.1 [2023-06-16 ucrt]).

## RESULTS

### Clinical Cohort Profile and Variant Distribution

Approximately 34% (244/721) of patients had a solid tumor primary diagnosis, ∼33% (240/721) had central nervous system (CNS) tumors, and the remaining ∼33% (237/721) had various hematolymphoid tumors (Supplementary Figure 1). After excluding duplicate germline variants and eliminating variants meeting BA1-stand-alone criteria, a total of 2376 unique variants were identified in our clinical cohort. These comprised 1288 missense, 660 silent, 309 splice-associated variants, 45 frameshift indels, 41 in-frame indels, 32 nonsense, and one variant in a non-coding gene (*TERC*). The 309 splice-associated variants comprised 11 canonical splice site variants and 298 in splicing regulatory regions extending from the last three bases of an exon to the first base of the following exon, not including the canonical splice site.

### Comparative Analysis of Variant Classification System

The analysis of variability in genetic variant classifications among ACMG/AMP 2015 and the point system reveals distinct distributions within the various variant categories (Figure 1a). Evaluation of overall pairwise concordance using Cohen’s Kappa statistic identified agreement between the ACMG/AMP 2015 guidelines and the point system classifications, with a Kappa coefficient of 0.63 (z = 49.7, p-value < 0.001; 95% CI: 0.59-0.68). The chi-squared test demonstrated a significant reduction in VUS when applying the point system by shifting the variant classification towards LB/B and LP/P (χ^2^ = 283.27, p-value < 0.001) (Supplementary Table 2).

**Figure 1.**
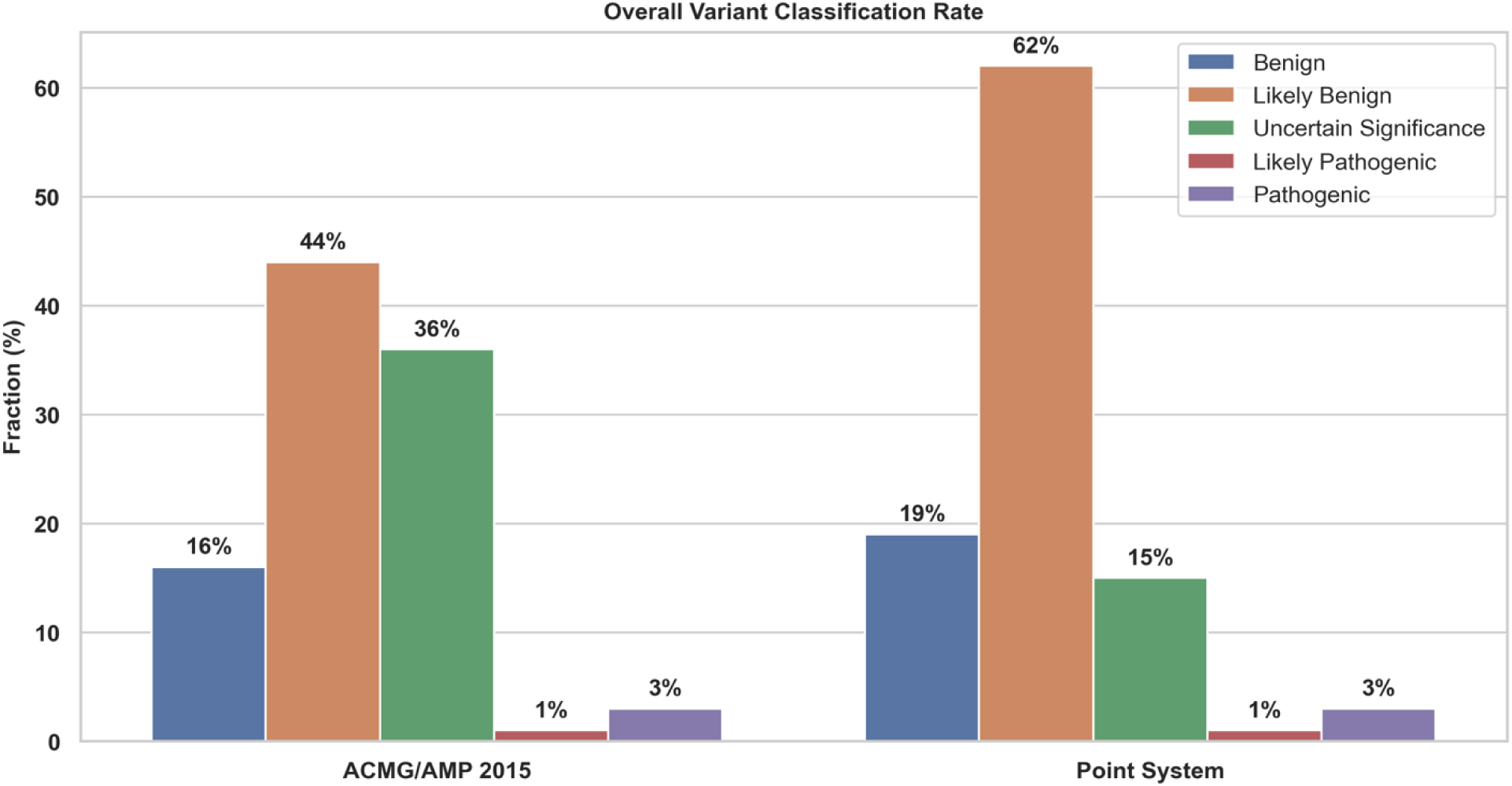
The overall variant classification rate across the ACMG/AMP 2015 and point system. The classification using strict ACMG/AMP 2015 guidelines versus point system depicts a ∼21% difference in the variant of uncertain significance rate (VUS), highlighting a level of agreement (Kappa coefficient of 0.63).

### Analysis of Discordant Results

To elucidate the main patterns of disagreement between the scoring systems and identify potential reasons for the decrease in VUS by the point system, an in-depth analysis was conducted on discordant calls between the ACMG/AMP 2015 guidelines and the point system. Among the 2376 unique variants analyzed, 23.5% (n=558) exhibited discordant classifications (Supplementary Table 3). These discordant variants were further divided into those that retained the originally defined evidence code strength from the ACMG/AMP 2015 guidelines (n= 444) and those that were assigned a modified evidence code strength (n = 114).

#### A) Variants with Original ACMG Evidence Codes

The predominant impact was on unique VUS, but it also affected unique P and LP variants (Figure 2). Among unique VUS called by the ACMG/AMP 2015, ∼18% (421/2376) were subsequently downgraded to LB, ∼0.8% (18/2376) were reclassified as B, and notably, one variant was upgraded to LP. Furthermore, two unique variants initially categorized as LP were upgraded to P. Conversely, the application of the point system resulted in two unique variants being downgraded from P to LP.

**Figure 2.**
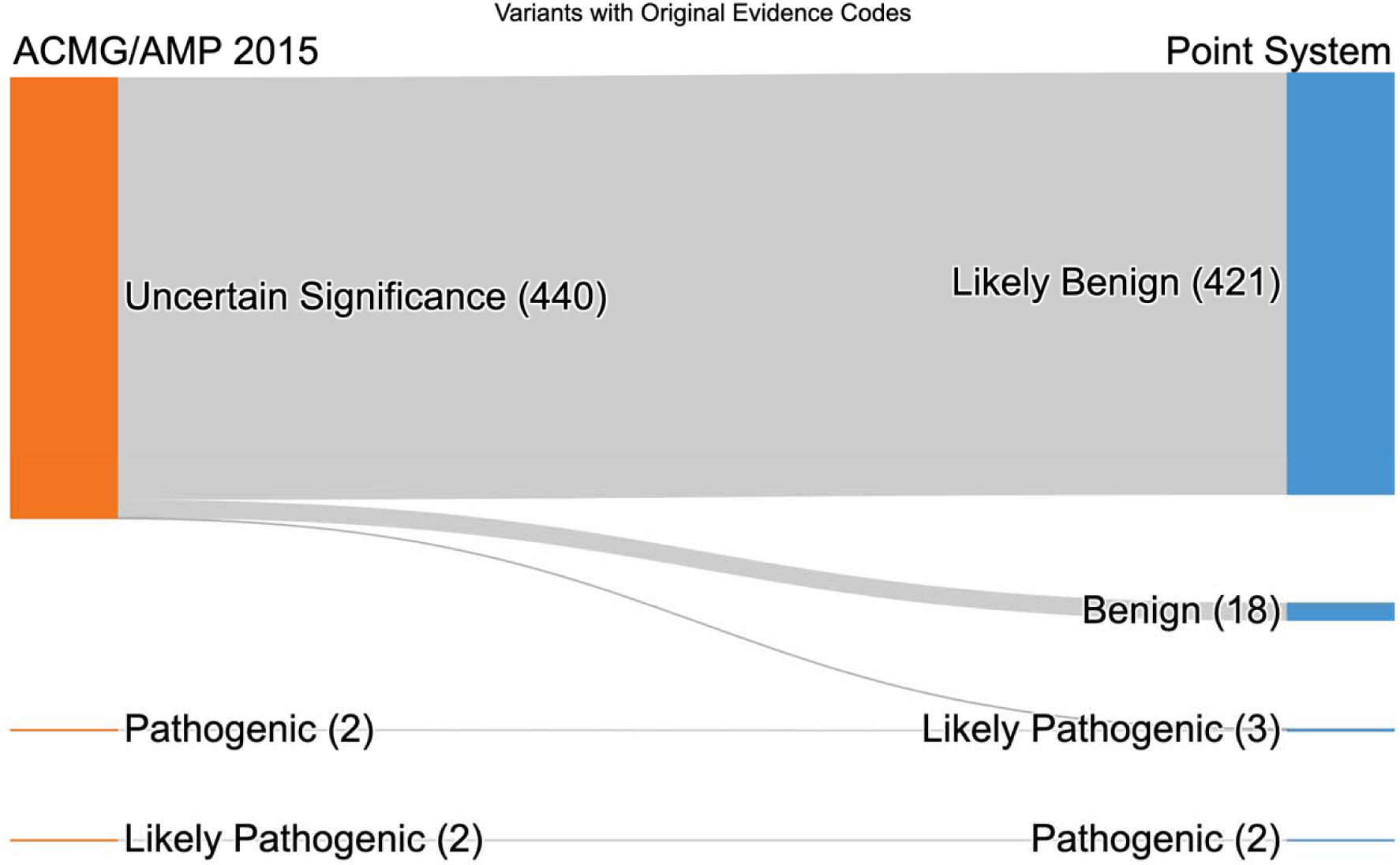
Sankey figure summarizing the classification changes between the ACMG/AMP 2015 guidelines and the point system. In variants classified using the original ACMG/AMP 2015 evidence code strengths (n=444), ∼18% (421/2376) of VUS were downgraded to LB using the point system, ∼0.8% (18/2376) were downgraded to B, and one variant was upgraded to LP. Two variants classified as LP were upgraded to P, and conversely, two variants were downgraded from P to LP.

Analysis of the evidence codes/categories, strengths, and corresponding point system scores within the discrepant categories (Supplementary Figure 2; Supplementary text) revealed 12% (288/2376) of unique variants with a score of -1, the minimum threshold in the point system for calling LB. The BP4 supporting evidence code was applied to most unique variants (283/2376), including 275 coding substitutions and eight intronic variants. Our application of PP3/BP4 criteria was dependent on the REVEL score, where a value ≤0.290 is criteria for BP4 (-1 point), ≥0.644 is criteria for PP3 (1 point), and 0.291-0.643 is inconclusive (0 points).^11^ One variant met criteria for BP2 alone and 4 variants met criteria for BP3 alone.

Approximately 6% (151/2376) of unique variants, classified as VUS according to ACMG/AMP 2015, had a score ≤ -2 using the point system. These variants were designated as VUS by ACMG/AMP 2015 guidelines because they did not meet a specific criterion and/or exhibited contradictory benign and pathogenic evidence. However, the variants achieved the minimum threshold score of -2 or -7 for classification as LB and B by the point system, respectively. Approximately 4% (98/2376) of unique variants were seen at score -4, driven by one benign strong evidence type (i.e., BS1, BS2, or BS3 each provides -4 points), with BS1 alone applied 87 times. The remaining ∼2% (53/2376) exhibited contradictory benign and pathogenic criteria. For example, one benign strong (-4 points) and one pathogenic supporting (1 point) criteria were frequently noted (n=20), resulting in a score of -3 (e.g., BS1 Strong with PP3 Supporting or PP2 Supporting) (Supplementary Figure 2; Supplementary text). The point system resulted in one unique variant, *CHEK2* NM_007194.4:c.190G>A (p.Glu64Lys), being upgraded from VUS to LP (score from 6 to 9 points). The *CHEK2* variant received PS3 Strong (4 points), PS4 Strong (4 points) and BP4 Supporting (-1 point). Additionally, two unique variants were upgraded from LP to P (score equal to or greater than 10 points). Those included *BLM* NM_000057.4:c.2250_2251insAAAT (p.Leu751LysfsTer25) and *POT1* NM_015450.2:c.1087C>T (p.Arg363Ter), where each variant received a PVS1 Very Strong (8 points) and one moderate evidence code (either PM3 or PM5 provides 2 points), resulting in a final score of 10 points. Conversely, due to a final score of 9 points, driven by two strong evidence codes (PS3 Strong and PS4 Strong) and one supporting evidence code (either PP1 Supporting or PP3 Supporting provides 1 point), the point system downgraded two unique variants, *MUTYH* NM_001128425.1:c.1187G>A (p.Gly396Asp) and *MITF* NM_198159.3:c.1255G>A (p.Glu419Lys), from P to LP (9 points).

#### B) Variants With Modified ACMG Evidence Codes

Approximately 5% (114/2376) of unique variants with discordant classifications between ACMG/AMP 2015 and the point system received at least one modified evidence code (Figure 3). The modified evidence code was either due to ClinGen recommendations or our internal committee/laboratory standard operating procedure (Supplementary Figure 3; Supplementary text). Approximately 3% (65/2376) of unique variants were classified as VUS by the ACMG/AMP 2015 guidelines due to contradictory criteria between benign and pathogenic evidence codes or evidence strength combinations not recognized in the original guidelines (e.g., PM3 Very Strong). Of these, 51 unique variants were downgraded to LB by the point system, including 28 unique variants with a score of -1 attained by two benign supporting evidence codes with one pathogenic supporting (e.g., BP4 Supporting; BP7 Supporting; PM2 Supporting) (Supplementary Figure 4a). The 14 unique VUS called by the ACMG/AMP 2015 included three that were upgraded to P and 11 variants that were upgraded to LP (Supplementary Figure 4b). PM3 Very Strong (8 points) evidence was applied to 4 unique variants based on the ClinGen guidance on variant phasing (https://clinicalgenome.org/working-groups/sequence-variant-interpretation/) and was combined with PVS1 Very Strong in three variants and therefore regarded as P. PM3 Very Strong was also combined with a pathogenic supporting evidence code (1 point) in one variant and therefore met the criteria for LP by the point system. The remaining upgraded LP variants comprised eight variants with PVS1 Very Strong with one pathogenic supporting evidence code; two variants were upgraded based on a combination of different pathogenic evidence codes that are not recognized by the ACMG/AMP 2015 guidelines.

**Figure 3.**
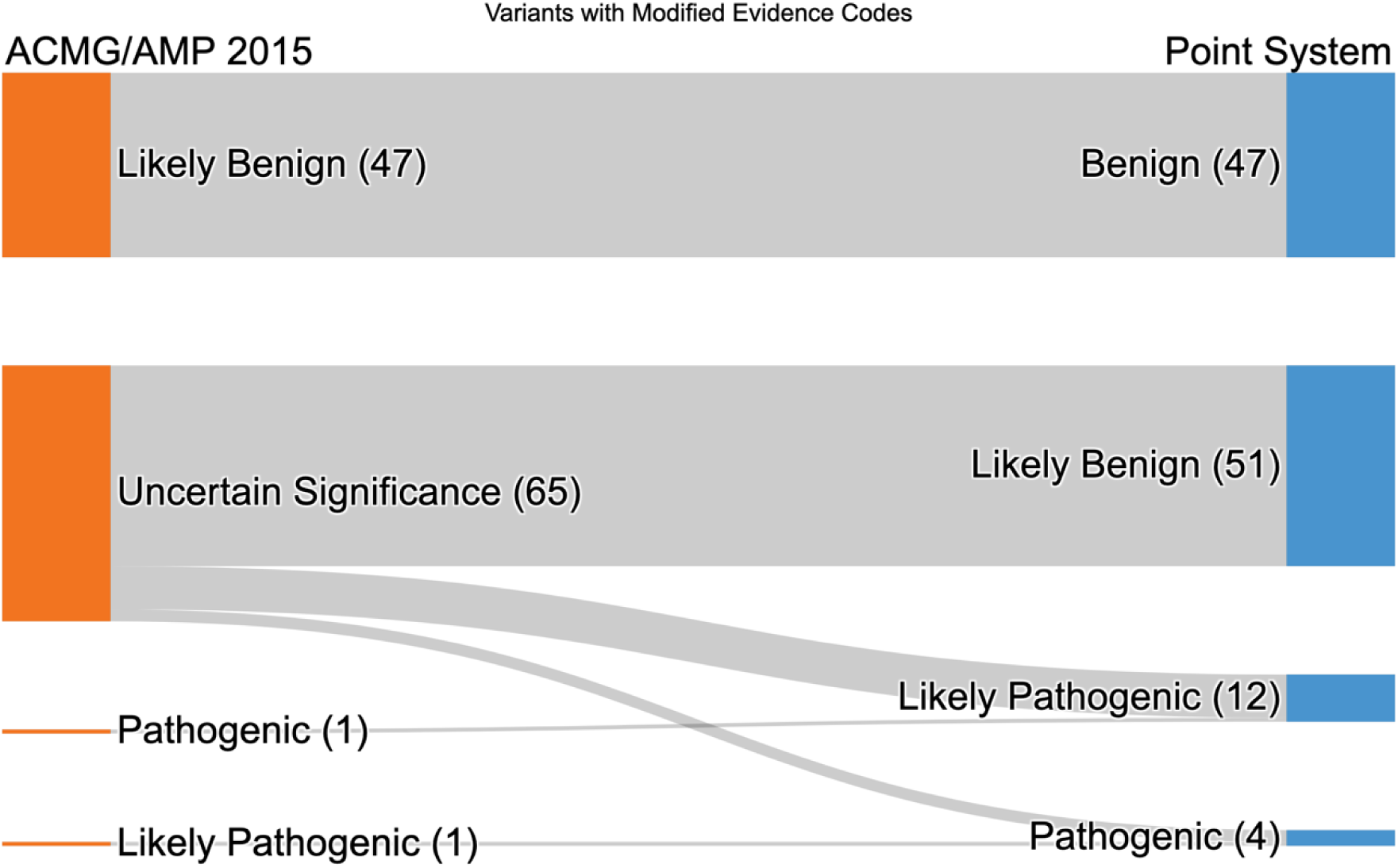
In variants with modified evidence code strengths (n=114), 2% (47/2376) of variants that were classified as LB using ACMG/AMP 2015 were downgraded to B, and 3% (65/2376) of VUS were reclassified as follows: 51 variants were downgraded to LB, 11 variants were upgraded to LP, and three variants were upgraded to P.

Interestingly, a variant in *BRCA2* NM_000059.3:c.9699_9702del that was classified as P by ACMG/AMP 2015 was subsequently downgraded to LP using the point system based upon the following evidence codes totaling 8 points: PS4 Strong; PVS1 Strong. The *BRCA2* variant is in the last exon and is not predicted to trigger nonsense-mediated-decay (NMD); therefore, PVS1 Very Strong was downgraded to PVS1 Strong according to the recommendations from Abou Tayoun and colleagues.^12^

Approximately 2% (47/2376) of unique variants with modified evidence codes applied that were categorized as LB based on ACMG/AMP 2015 were further downgraded to B by the point system. With the exception of BA1, the original ACMG/AMP 2015 recommendations typically required ≥ 2 strong evidence codes for benign and either one strong and one supporting evidence code or ≥ 2 supporting evidence codes for LB. In total, 45 unique variants with a score of -7 were downgraded from LB to B using the point system (Supplementary Figure 4c).

### VUS Sub-tiering Using the Point System

A recent study by Rehm and colleagues proposed sub-tiering VUS into VUS-Low, VUS-Mid, and VUS-High.^13^ To further assess the utility of the point system in sub-tiering VUS, we examined the point system score and variant type for each unique VUS classified by the point system (Figure 4; Supplementary Table 3). The VUS were categorized as follows: VUS-Low for scores of 0 and 1, VUS-Mid for scores of 2 and 3, and VUS-High for scores of 4 and 5. Of the 354 unique VUS called by the point system, ∼11.5% (274/2376) were regarded as VUS-Low, ∼3% (72/2376) were VUS-Mid, and less than 1% (8/2376) were classified as VUS-High.

**Figure 4.**
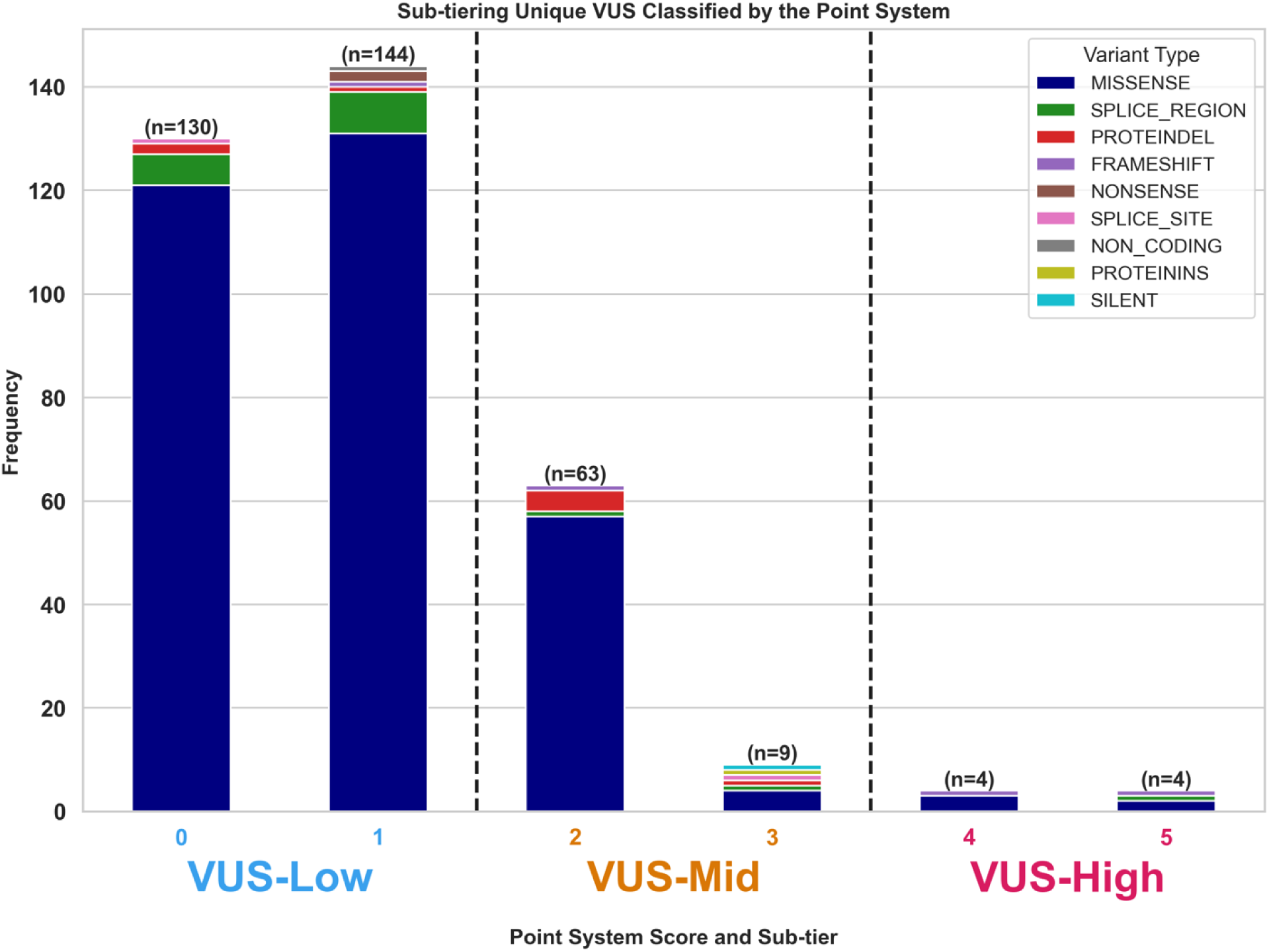
Sub-tiering the unique VUS classified by the point system. Dashed lines delineate the unique VUS with variants binned into VUS-Low (0-1 points); VUS-Mid (2-3 points) and VUS-High (4-5 points). ∼77% of unique VUS were in the VUS-Low category indicating that they have little evidence available to facilitate classification.

Among the 354 unique VUS called by the point system, missense variants were the most prevalent (n=318). Of these, 298 (94%) were supported only by population data from the gnomAD v2.1.1 database (PM2 Supporting) and/or bioinformatic prediction data (BP4, PP3 or PP2; Supplementary text). In comparison with the ACMG/AMP 2015, there were 751 missense variants classified as of uncertain significance. Of these, 612 (81%) only had population data obtained from the gnomAD v2.1.1 database (BS1, BS2, PM2) and/or bioinformatic prediction data (PP3, BP4, PP2).

## DISCUSSION

We compared the scoring rules for the qualitative ACMG/AMP 2015 guidelines and the quantitative Bayesian point-based system and their respective effects on germline variant classification. Our analysis highlighted that the point system reduces the unique number of VUS by either downgrading them to LB/B or upgrading them to LP/P. We observed a discrepancy in variant scoring between the two methodologies in ∼23.5% of unique variants analyzed. Among discrepant variants, ∼21% were attributed to a decrease in unique VUS rates (Figure 5) that was driven by three primary modifications: 1) ∼12% reduction in unique VUS with a point system score of -1, primarily due to a single benign supporting evidence code (most notably BP4 Supporting in 283 variants); 2) ∼4% reduction in unique VUS with a point system score of -4 linked to a single benign strong evidence code (e.g., BS1, BS2, or BS3 in 98 variants); and 3) ∼5% reduction in unique VUS with conflicting evidence codes or criteria not acknowledged by the current ACMG/AMP 2015 guidelines. The remaining ∼2.5% of discrepant unique variants were related to the reclassification from LB to B by the point system and, to a lesser extent, changes between LP and P classifications.

**Figure 5.**
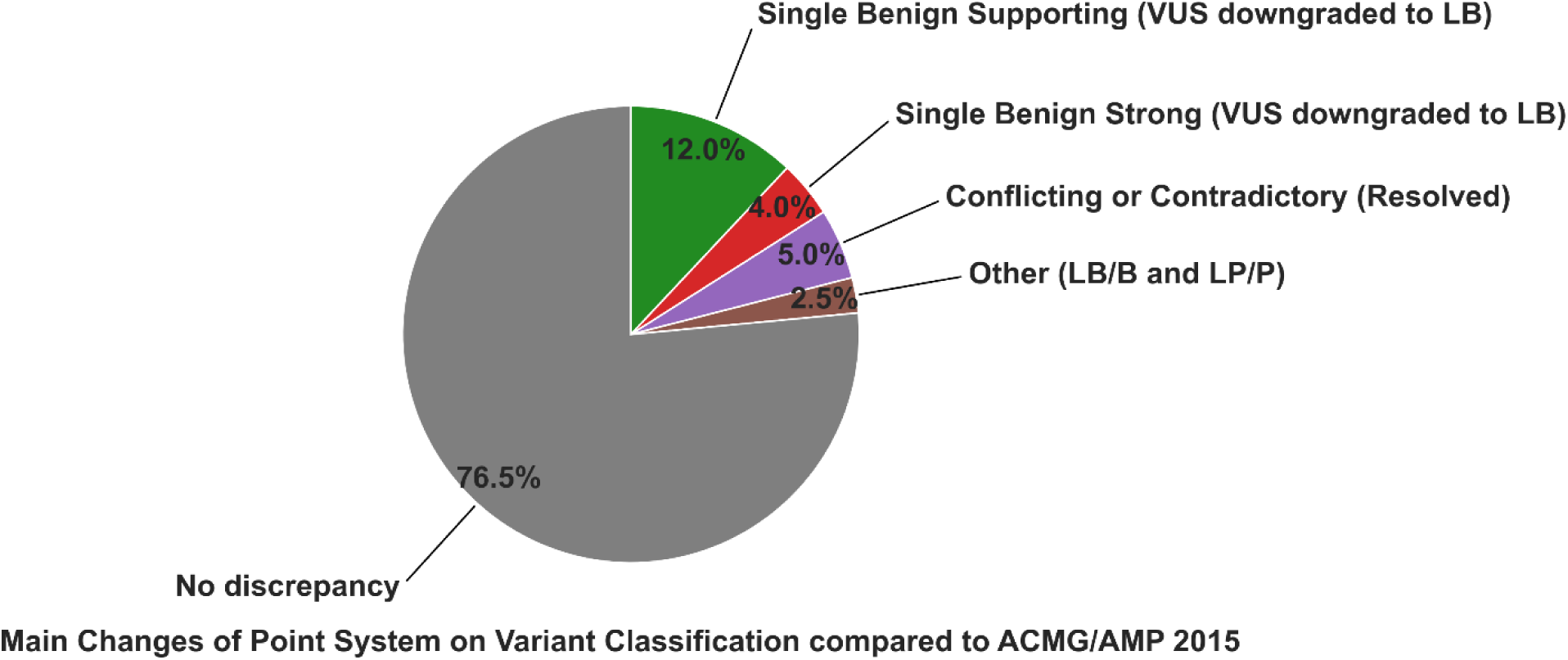
Main changes to variant classification using the point system compared to ACMG/AMP 2015 guidelines. A discrepancy between classifications derived from the point system and the ACMG/AMP 2015 guidelines was observed for 23.5% of the unique variants in our dataset. Among these discrepancies, ∼21% were attributed to a decrease in the number of VUS, primarily due to ∼12% downgraded to LB driven by single benign supporting evidence (mostly BP4 Supporting), ∼4% downgraded to LB driven by single benign strong evidence (mostly BS1 Strong), and ∼5% changed from resolving conflicting/contradictory evidence or criteria not recognized by the current ACMG/AMP 2015 guidelines. The remaining ∼2.5% of differences are related to changes between LB and B or LP and P classifications.

To elucidate the primary causes of the discrepancies observed between these two classification systems, it is essential to consider the seminal insights from the research undertaken by Tavtigian and colleagues.^7^ Tavtigian and colleagues uncovered a distinct pattern within the ACMG/AMP 2015 criteria where one criterion from a stronger evidence category could equivalently be replaced by two criteria from the next lower strength category or by four criteria from two subsequently lower categories. In the ACMG/AMP 2015, an LB classification can be achieved when a variant has at least two supporting benign evidence types; within the point system, one strong piece of benign evidence (e.g., BS1-4) would be equivalent to four pieces of supporting benign evidence (BP1-BP7), thereby positing that a single strong evidence type could suffice to reclassify a variant from VUS to LB. Second, inference of the point-system scale derived from a Prior_P of 0.1 resulted in a single piece of benign evidence or equivalent conflicting evidence with a point score of -1 being sufficient to downgrade a VUS to LB.^8^ This critical difference from the criteria set forth in the ACMG/AMP 2015 guidelines explains a ∼12% reduction in the unique VUS rate by the point system. The ACMG/AMP 2015 guidelines recommend that experts should use their judgment when encountering conflicting/contradictory evidence codes; however, the ability of the point system to address these conflicts suggests a path for large-scale automated classification of variants.^1,7^ For instance, using the point system, a combination of BS1 Strong with PP3 Supporting evidence would result in a net score of -3 (LB), demonstrating that one pathogenic supporting evidence type is unable to negate the effect of one benign strong evidence. The point system also aids in resolving criteria not directly recognized by the ACMG/AMP 2015. For instance, a single very strong evidence type alone or in combination with a supporting one could result in an LP designation. Combinations like BP4, BP7, and PM2 Supporting classified as LB (-1 point) and VUS (0 point) with PM2 Moderate.

Another key advantage of the point system is that it provides a structured method for categorizing VUS into sub-tiers and facilitates the prioritization of variants for reanalysis. In the study by Rehm and colleagues, a subset of clinical laboratories subclassified VUS as VUS-High, VUS-Mid, or VUS-Low, with some laboratories opting to report only VUS that have stronger evidence (e.g., VUS-High), which seemed to be favored by clinical providers.^13^ Utilization of the point system could facilitate a more standardized approach to sub-tiering VUS based on their scores and variant types. In our dataset, approximately 15% of the unique VUS identified by the point system were deemed VUS-Low, representing ∼11.5% (274/2376) of unique variants, followed by ∼3% deemed VUS-Mid. Conversely, only 8 unique variants, less than 1%, were classified as VUS-High. The VUS-High can be prioritized by clinical laboratories for periodic reanalysis and could assist clinicians in guiding management and determining the need for clinical follow-up.^13^

Future large-scale prospective studies are necessary to more definitively assess the validity and efficacy of the point system in variant classification. However, preliminary evidence from our study suggests that the point system can significantly reduce the VUS rate through the application of a single benign strong evidence code and by helping to resolve conflicting/contradictory evidence types. This is further supported by the fact that several ClinGen variant curation expert panel working groups (e.g., *APC, CDH1,* etc.) apply BS1 strong alone with a certain minor allele frequency cut-off to classify a variant as LB – a change that closely aligns with the point system scoring method.^14,15^ It is important, however, to exercise caution when downgrading VUS to LB based on single benign supporting evidence codes or equivalent conflicting evidence with a point score of -1. Tavtigian and colleagues have raised the argument that the point threshold for classifying a variant as LB could be set at −2, aligning it with the ACMG/AMP 2015 guidelines, rather than relying on the threshold set by a prior probability of <0.10.^8^ In addition, the application of BP4 based on *in-silico* predictions algorithms may lead to false negatives in the evaluation of gain-of-function missense variants, as reported in *SAMD9* and subset of genes in mTOR pathway.^16,17^ Lastly, it would be essential to rule out possible splicing impacts of substitutions/missense variants using splicing prediction algorithms to appropriately apply BP4 as stand-alone evidence for an LB classification. This underscores the potential necessity to refine the application of single benign supporting evidence within the point system, particularly in relation to *in-silico* predictions. An alternative strategy might involve expanding the VUS-Low category to encompass variants scored as -1, 0, and 1, offering a more nuanced approach to circumvent this ambiguity.

Reporting VUS can lead to either over- or under-management, with the potential for unnecessary interventions or missed early detection opportunities, respectively.^13^ Additionally, failing to report impactful VUS can result in overlooked diagnoses and/or delays in screening and treatment. Periodic reanalysis frequently leads to the reclassification of VUS, with around 80-90% being downgraded to LB/B, while about 10-20% are upgraded to LP/P.^18,19^ The National Comprehensive Cancer Network (NCCN) guidelines recommend not altering patient management in individuals with VUS but advise screening based on personal and family histories.^20^ It is, therefore, crucial to carefully balance and optimize VUS reporting to prevent the potential negative consequences of under-reporting or mismanaging VUS.^13^

Our analysis has several limitations and we recognize the potential for variability in the classification of variants and the application of evidence types across different clinical laboratories, as highlighted by the literature.^2^ To mitigate this, our approach involved a direct comparison of the ACMG/AMP 2015 guidelines and the point system, further stratifying the data by original evidence codes and modified ones. Our analysis concentrated on genes associated with hereditary cancer syndromes and did not address the influence of the point system on genes related to Mendelian disorders that are not associated with cancer predisposition syndromes or somatic variant curation.^21^ Subsequent large-scale studies will be critical for validating our findings and for discerning additional levels of agreement or disparity between the two models.

In conclusion, our study underscores the need to continually refine the scoring systems used in genetic variant classification. By comparing the ACMG/AMP 2015 guidelines against the point system, our study demonstrates that the point system would have a substantial impact in reducing the VUS rate and could facilitate sub-tiering in clinical reports. Future large-scale prospective studies to evaluate incorporating the VUS sub-tiering with patient’s personal/family history are warranted, which could enable healthcare providers to offer more precise and tailored screening guidelines and avoid unnecessary interventions.

## Data Availability

All of the relevant data used in this study has been provided in the tables, or relevant supplementary information.

## Web Resources

ClinVar, https://www.ncbi.nlm.nih.gov/clinvar/

Genome Aggregation Database (gnomAD), https://gnomad.broadinstitute.org/

REVEL, https://sites.google.com/site/revelgenomics/

ClinGen: https://clinicalgenome.org/working-groups/sequence-variant-interpretation/

Criteria Specification Registry, https://cspec.genome.network/

The National Comprehensive Cancer Network (NCCN), https://www.nccn.org

## Acknowledgments

We thank all the patients and their families at St. Jude Children’s Research Hospital (SJCRH) for their contribution to the biological specimens used in this study.

## Author Contributions

Conceptualization: M.K.E.; Data curation: M.K.E., J.L.M., P.R.B., T.C., V.P.; Formal analysis: M.K.E., J.L.M., P.R.B., T.C., V.P.; Supervision: M.K.E., J.L.M., P.R.B.; Statistical analysis: L.T., S.S.M.; Writing-original draft: M.K.E., J.L.M., P.R.B.,; Writing-review & editing: all authors.

## Ethics Declaration

The study was reviewed by the St. Jude Children’s Research Hospital Institutional Review Board, which determined that this project did not meet the criteria for human subjects research and, therefore, did not require full institutional review board approval. All patients consented to clinical analysis and individual reporting of germline sequencing data.

