## Supplementary Figures for "Evaluation of Bayesian Point-Based System on the Variant Classification of Hereditary Cancer Predisposition Genes"

### Slide 1
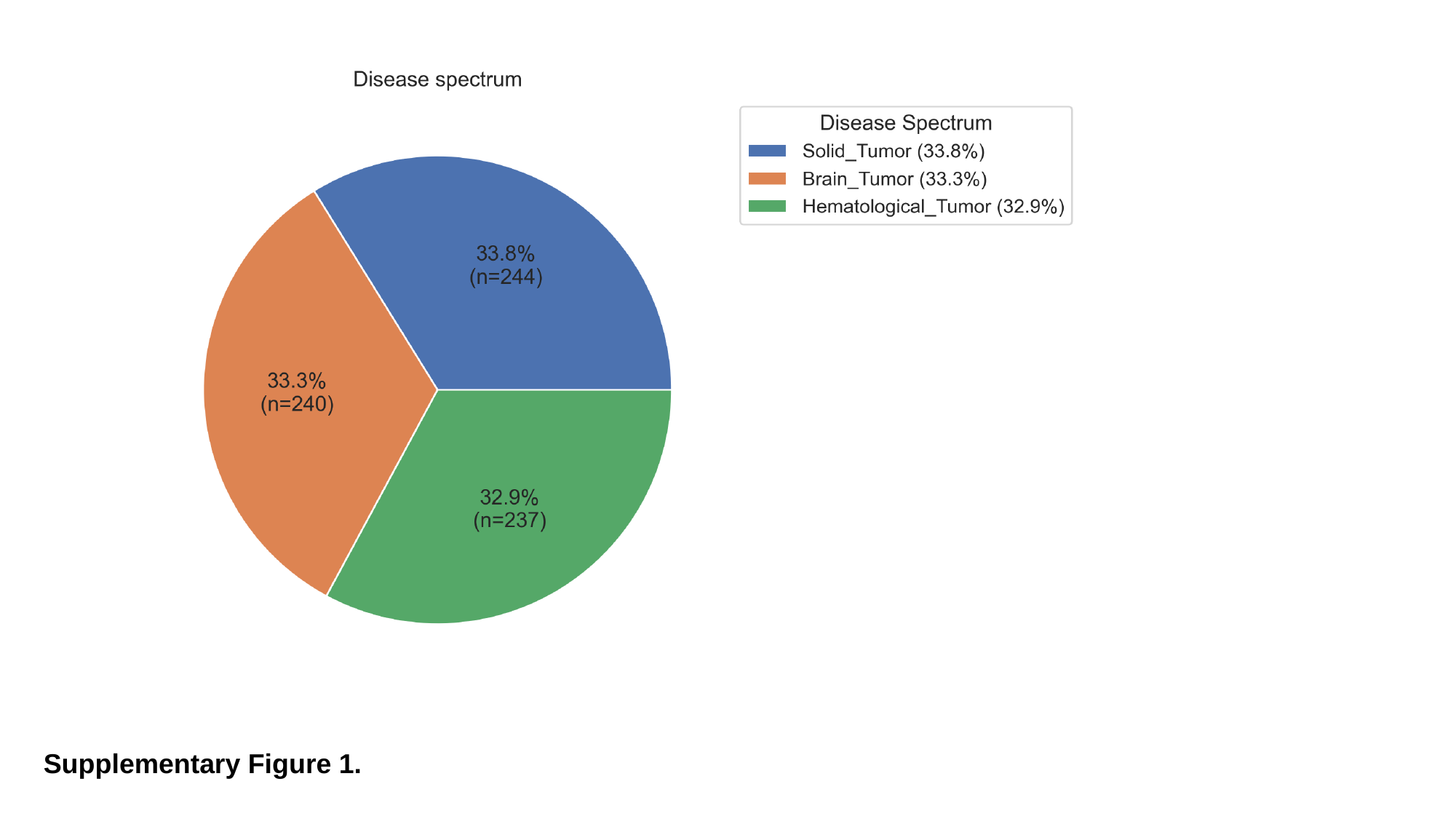

Supplementary Figure 1.

### Slide 2
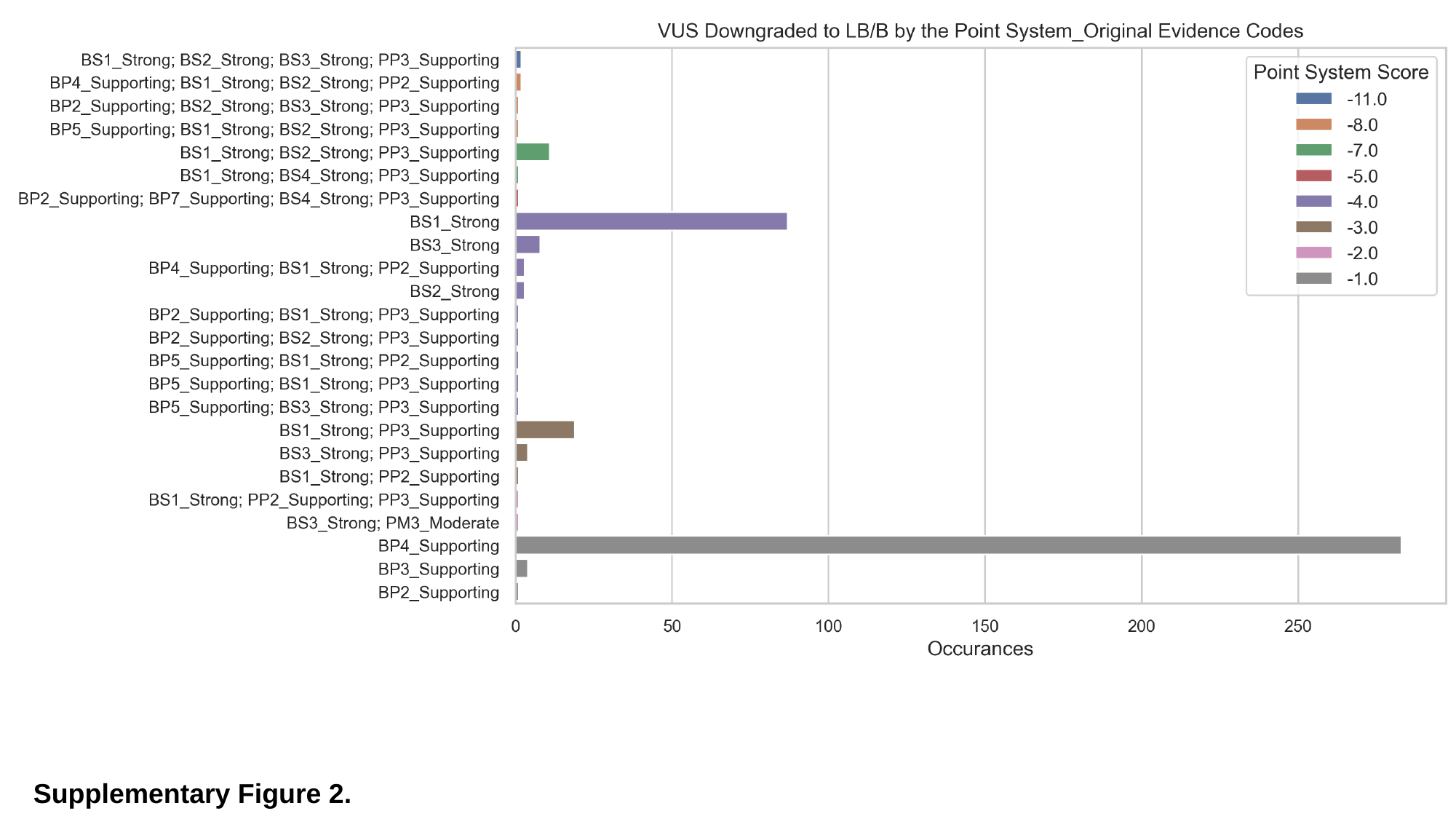

Supplementary Figure 2.

### Slide 3
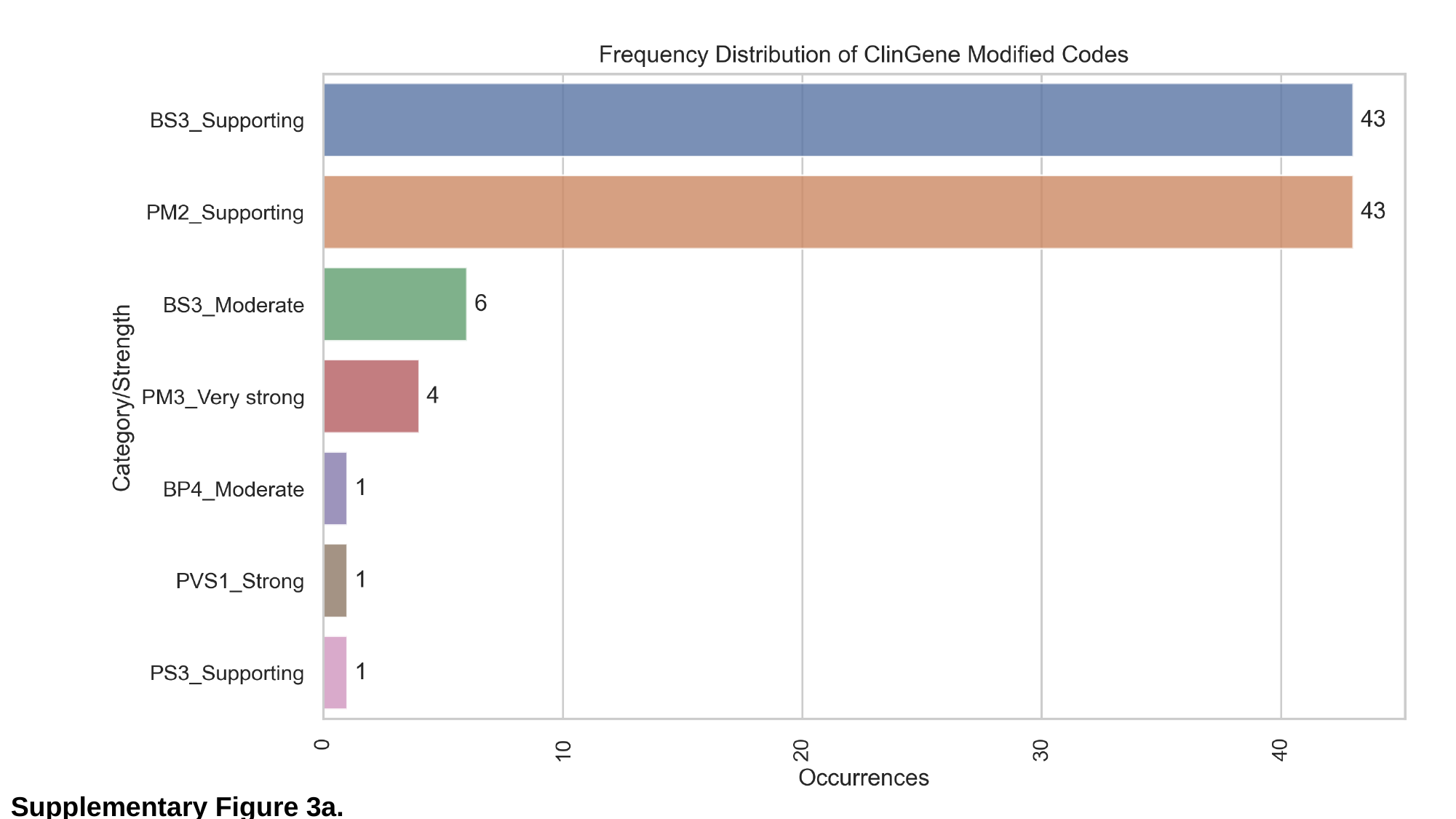

Supplementary Figure 3a.

### Slide 4
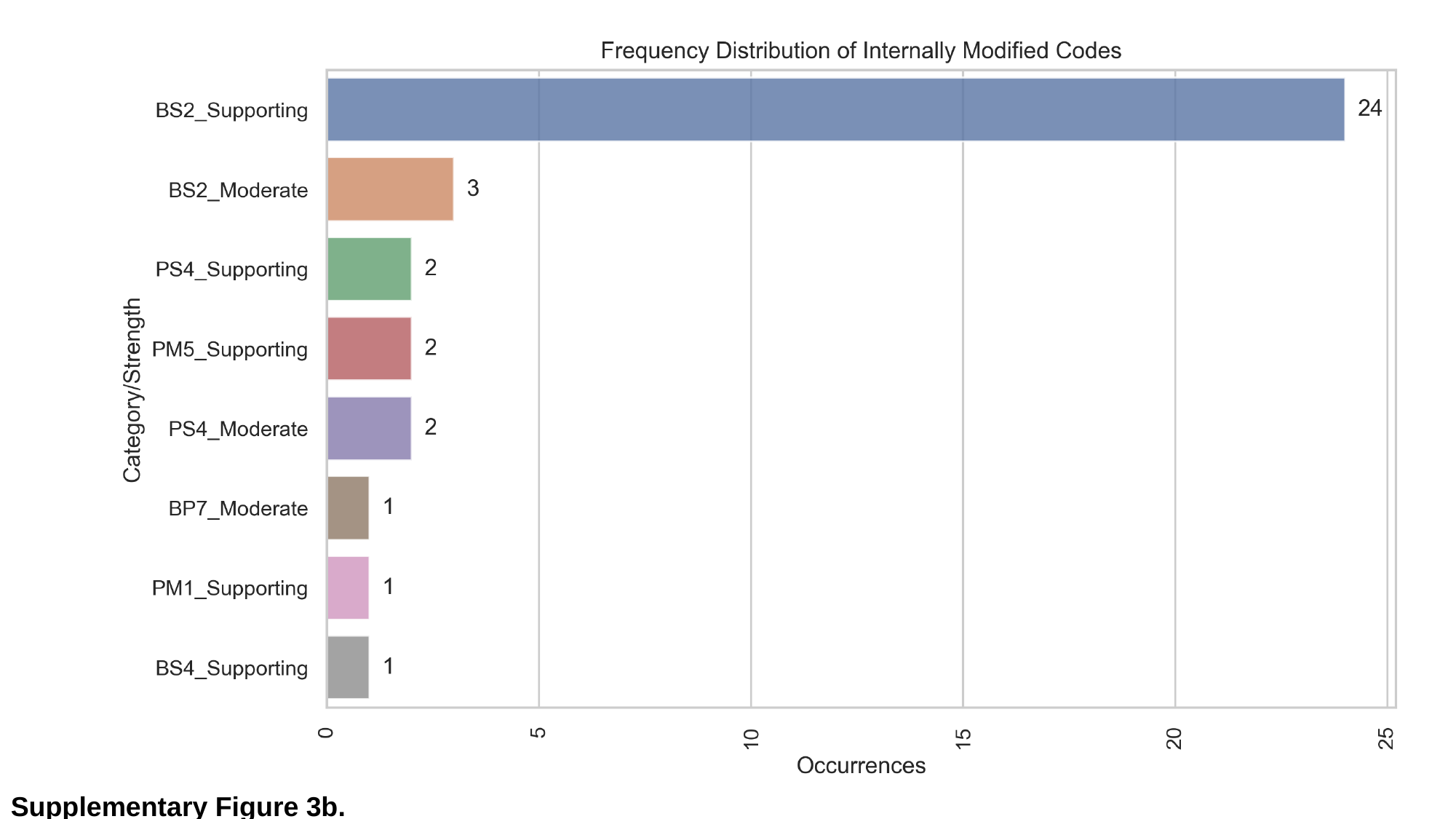

Supplementary Figure 3b.

### Slide 5
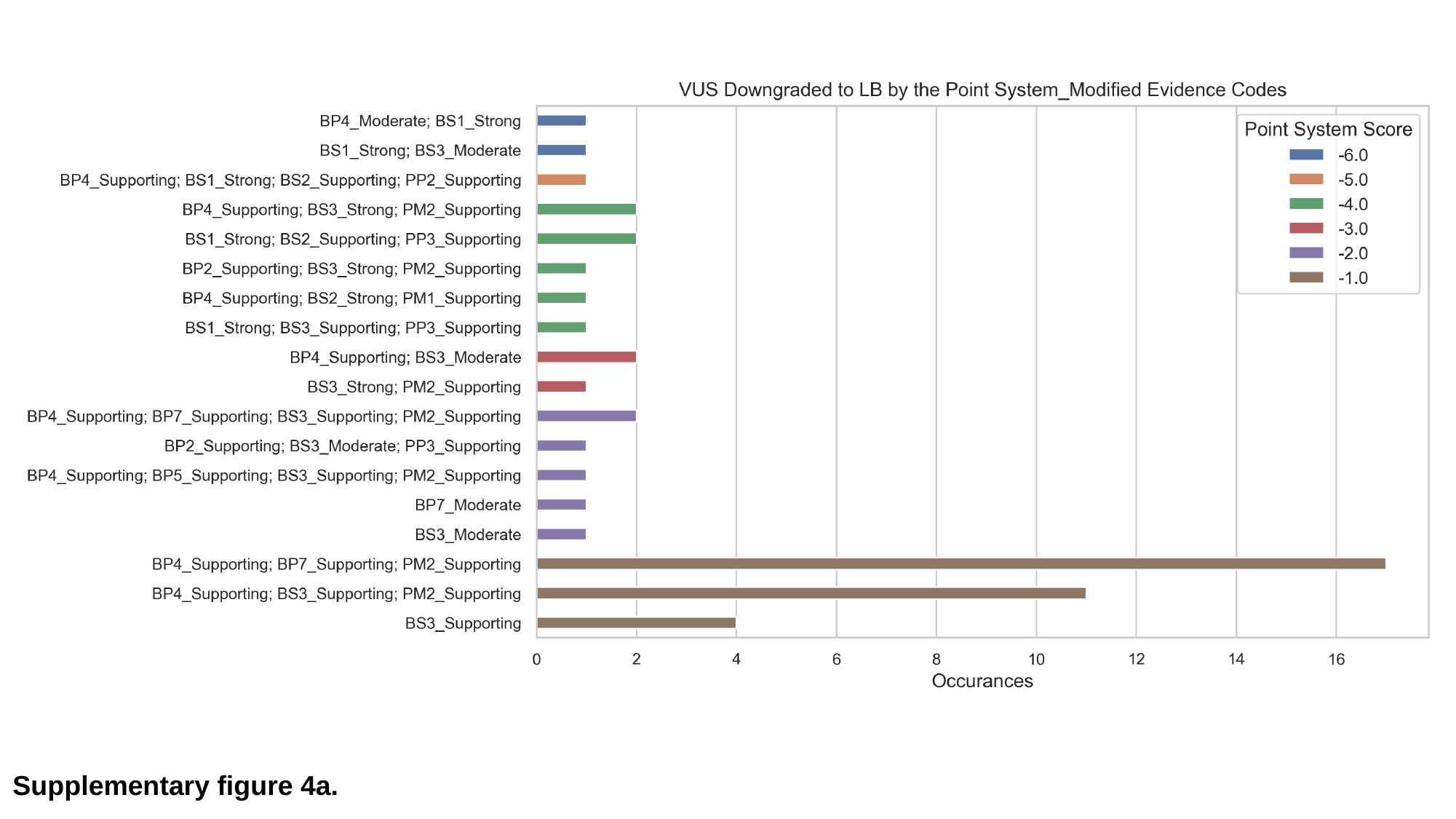

Supplementary figure 4a.

### Slide 6
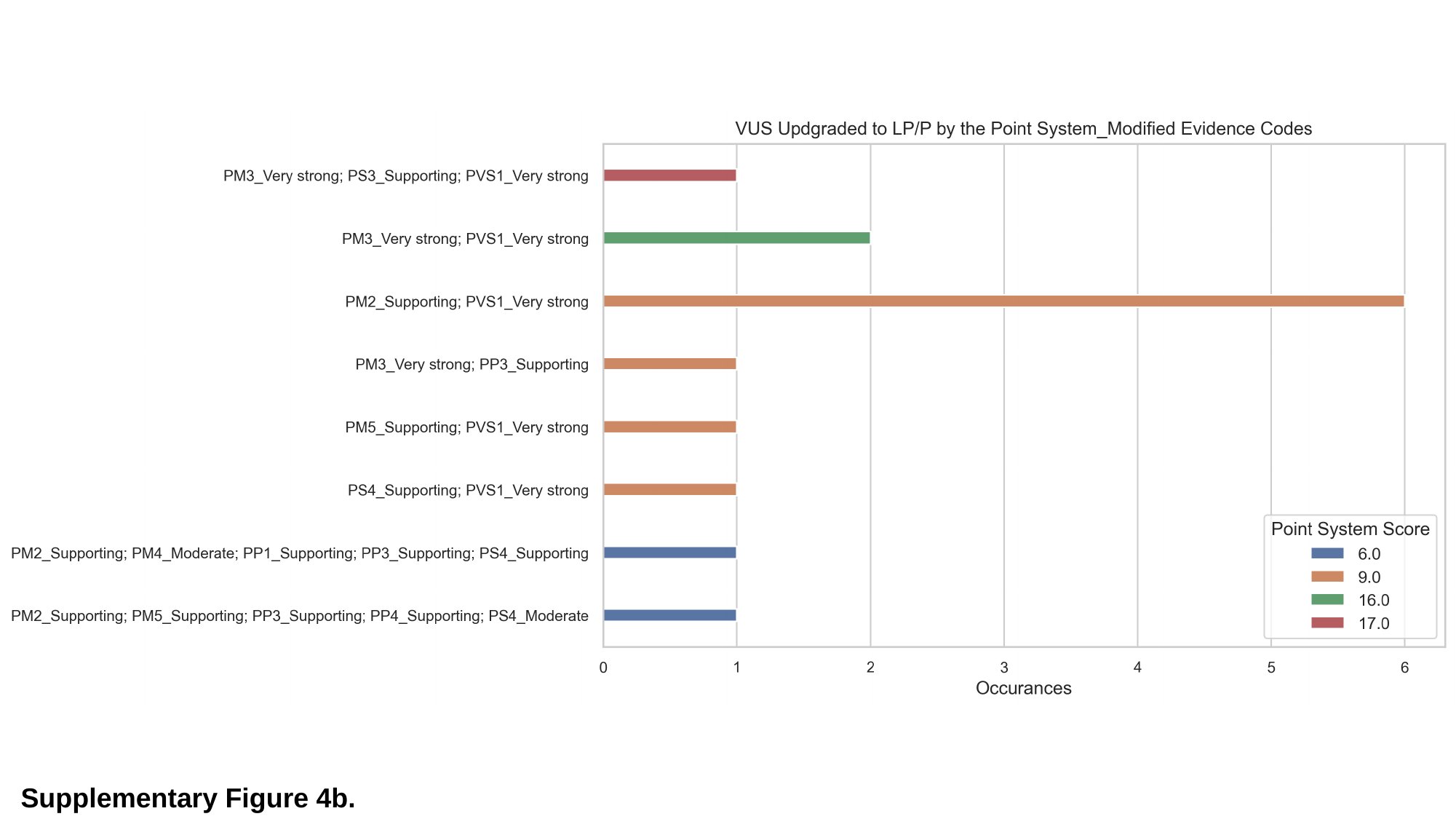

Supplementary Figure 4b.

### Slide 7
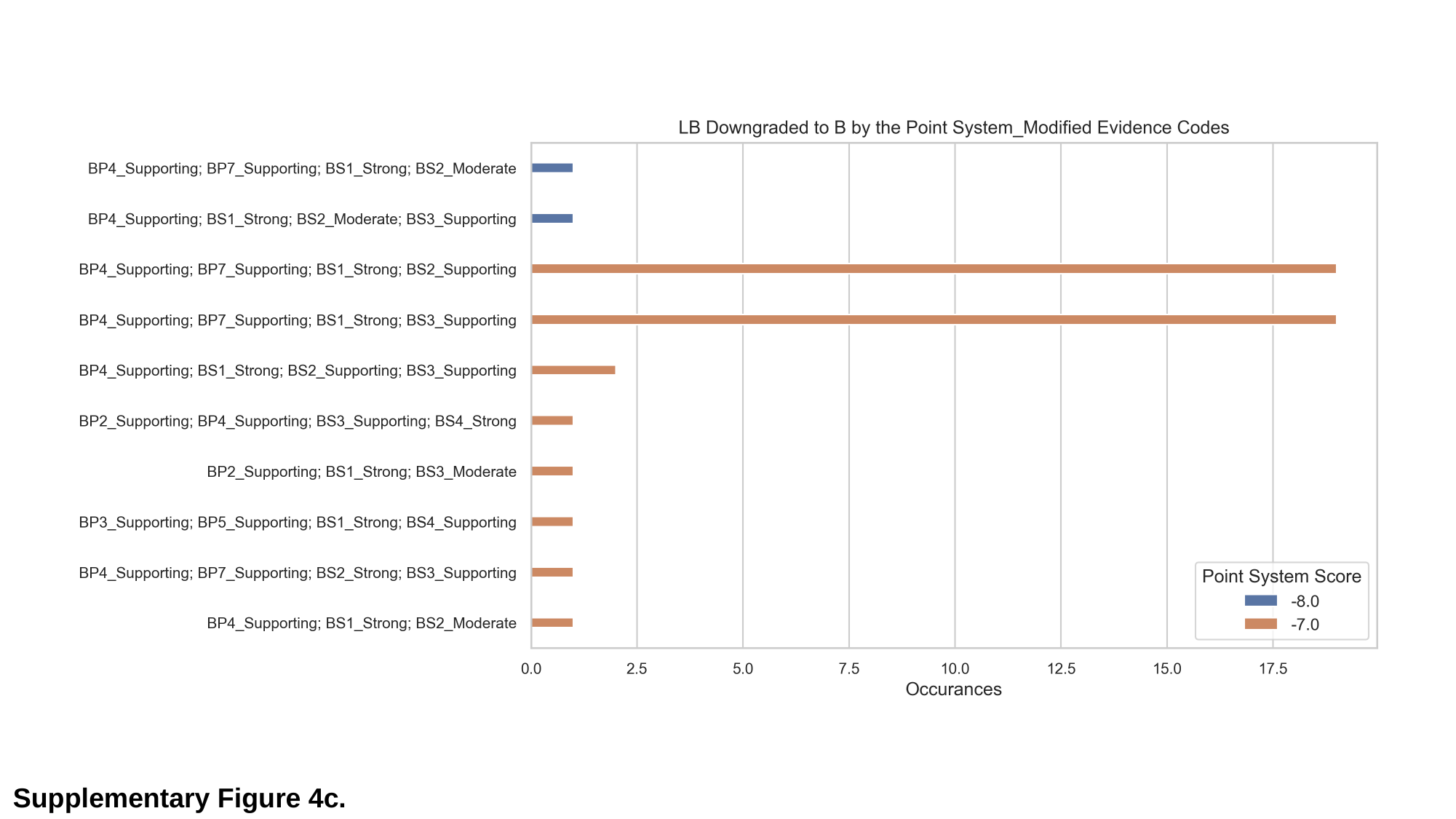

Supplementary Figure 4c.
