## Supplementary Text for "Evaluation of Bayesian Point-Based System on the Variant Classification of Hereditary Cancer Predisposition Genes"

A) **Variants with Original ACMG Evidence Codes (Supplementary text)**

The evidence code of BP4 Supporting was applied to 12% (283/2376) of unique variants. Two variants had a score of -2 and 24 variants had a score of -3 characterized by conflicting benign and pathogenic evidence codes (e.g., BS1 Strong; PP3 Supporting). Four variants had *in silico* evidence predicting pathogenicity (PP3 Supporting) that conflicted with published functional studies (BS3 Strong). This included two variants in *MSH2*, one variant in *BRCA1*, and one variant in *BRCA2*. The *MSH2* NM_000251.3:c.728G>A (p.Arg243Gln) variant has a REVEL score of 0.84 and was shown to function similar to wild-type in a massively parallel screen and in a methylation tolerance-based functional assay.^1,2^ The *MSH2* NM_000251.2:c.1730T>C (p.Ile577Thr) variant has a REVEL score of 0.928 and was shown to be functionally neutral in a massively parallel screen.^1^ The *BRCA1* NM_007294.4:c.5317A>T (p.Thr1773Ser) variant has a REVEL score of 0.682 and was shown to be functionally neutral in a saturation genome editing assay.^3^ The *BRCA2* NM_000059.3:c.7565C>T (p.Ser2522Phe) variant has a REVEL score of 0.713 and was shown to be functionally neutral in a homology-directed DNA repair assay.^4^

Approximately 4.4% (106/2376) with a score of -4, including 98 variants with one benign strong evidence type (BS1, BS2 or BS3) and 8 variants with benign strong and supporting evidence type with one pathogenic supporting evidence. BS1 Strong evidence was able to downgrade VUS to LB in 87 variants. 16% (393/2376) of these LB variants were missense variants, 12 were splice region variants, 5 had silent changes, 8 were in-frame indels, 7 were frameshift variants, and one was a splice site change that received strong evidence (-4). One silent variant with a score of -5. In our dataset, 18 missense variants were downgraded from VUS to B by the point system and were characterized by conflicting evidence with the enrichment of benign evidence types together with one pathogenic supporting evidence type.

B) **Variants with Modified ACMG evidence codes**

The most common modified evidence codes based on ClinGen recommendations were BS3 Supporting and PM2 Supporting, with 43 instances each (Supplementary Figure 3a). BS3 Moderate and PM3 Very strong were noted with 6 and 4 counts, respectively. Other categories, BP4 Moderate, PVS1 Strong, and PS3 Supporting, are the least represented, each with only a single occurrence. Our committee review's most common modified evidence codes were BS2 Supporting, featuring 24 occurrences. Other categories were also noted (Supplementary Figure 3b)

Fifty-one variants were downgraded in this modified evidence category from VUS to LB either due to contradictory criteria between benign and pathogenic evidence codes or due to certain combinations that are not recognized in the ACMG/AMP 2015 guidelines (e.g., BP4 Moderate and BS1 Strong). A total of 32 variants reached the minimum cutoff of likely benign (-1) and included 28 downgraded with two benign supporting evidence codes with one pathogenic supporting: BP4 Supporting, BP7 Supporting, PM2 Supporting was seen in 11 silent variants, and 6 splice region/intronic variants. The combination of BP4 Supporting, BS3 Supporting, and PM2 Supporting was applied to 10 splice region variants and 1 missense variant. 4 missense variants received BS3 Supporting. The remaining 19 variants (14 missense, 2 splice intronic regions, two silent and one inframe alteration) downgraded by the point system reached the point system score of equal to or less than -2.

The predominant patterns (n=38) in LB variants downgraded to B included BP4 Supporting, BP7 Supporting, BS1 Strong with either BS2 Supporting or BS3 Supporting. Other combinations were also seen and only two variants have had a score of -8.

**VUS Sub-tiering Using the Point System**

Analysis of the distribution point system scores of unique missense variants revealed a high frequency of variants with scores of 0 and 1 (121 and 131 occurrences, respectively), but with a notable decrease of variants with higher scores indicating more evidence criteria met; 57 variants had a score of 2, and 9 total variants had scores ranging from 3 to 5 (Supplementary Table 3b). For unique missense VUS with a point system score of 0, both a pathogenic and benign supporting evidence type were applied, leading to a net score of 0, with the most commonly occurring combinations being BP4 Supporting with PM2 Supporting (n=101) and BP4 Supporting with PP2 Supporting (n=13). The majority of unique missense VUS with a score of 1 harbored a single pathogenic supporting evidence code (e.g., PP3 Supporting, n = 69; PM2 Supporting, n = 41). Variants with a score of 2 were frequently associated with two pathogenic supporting evidence codes (e.g., PM2 Supporting; PP3 Supporting, n =43). **Nine missense** VUS with scores of 3 and above had either several pathogenic supporting evidence codes or evidence codes with moderate or strong strength.
